# Comparisons of new-onset peripheral arterial disease in Type 2 diabetes mellitus patients exposed to SGLT2I, DPP4I or GLP1a: a population-based cohort study

**DOI:** 10.1101/2023.08.21.23294389

**Authors:** Oscar Hou-In Chou, Zhiyao Luo, Cheuk To Skylar Chung, Jeffrey Chan, Huixian Li, Ishan Lakhani, Sharen Lee, Qingpeng Zhang, Tong Liu, Wing Tak Wong, Bernard Man Yung Cheung, Gregory Y. H. Lip, Gary Tse, Fung Ping Leung, Jiandong Zhou

## Abstract

**Background:** Sodium-glucose cotransporter-2 inhibitors (SGLT2I) have been suggested to have beneficial effects against atherosclerotic cardiovascular disease. The comparative risks of new onset peripheral arterial disease (PAD) between SGLT2Is, dipeptidyl peptidase-4 inhibitors (DPP4Is) and glucagon-like peptide-1 receptor agonist (GLP1a) remain unknown.

**Objective:** This real-world study aims to compare the risks of PAD upon exposure to SGLT2I and dipeptidyl peptidase-4 inhibitors (DPP4I).

**Methods:** This was a retrospective population-based cohort study of patients with type-2 diabetes mellitus (T2DM) on either SGLT2I or DPP4I between 1st January 2015 and 31st December 2020 using a territory-wide registry in Hong Kong. The primary outcome was new-onset PAD. The secondary outcome was all-cause mortality. Propensity score matching (1:1 ratio) using the nearest neighbour search was performed. Multivariable Cox regression was applied to identify significant associations. A three-arm sensitivity analysis including the GLP1a cohort was conducted.

**Results:** This cohort included 75470 T2DM patients (median age: 62.3 years old [SD: 12.8]; 55.79 % males). The SGLT2I and DPP4I groups consisted of 28753 patients and 46717 patients, respectively. After matching, 186 and 256 patients suffered from PAD in the SGLT2I and DPP4I groups respectively, over a median follow-up of 5.6 years. SGLT2I use was associated with lower risks of PAD (Hazard ratio [HR]: 0.85; 95% Confidence Interval [CI]: 0.67-0.98) compared to DPP4I use after adjustments for demographics, comorbidities, medications, renal function, and diabetic laboratory tests. Similar associations were observed in subgroup analyses in male patients above 65 years old, with hypertension, and low HbA1c levels. In the sensitivity analysis, SGLT2I was not associated with lower risks of PAD compared to GLP1a (HR: 0.88; 95% CI: 0.65-1.18). The results remained consistent in the competing risk and the sensitivity analyses.

**Conclusions:** SGLT2I use amongst T2DM patients was associated with lower risks of new-onset PAD and PAD-related outcomes when compared to DPP4I after adjustments.

**Illustrated Abstract:** 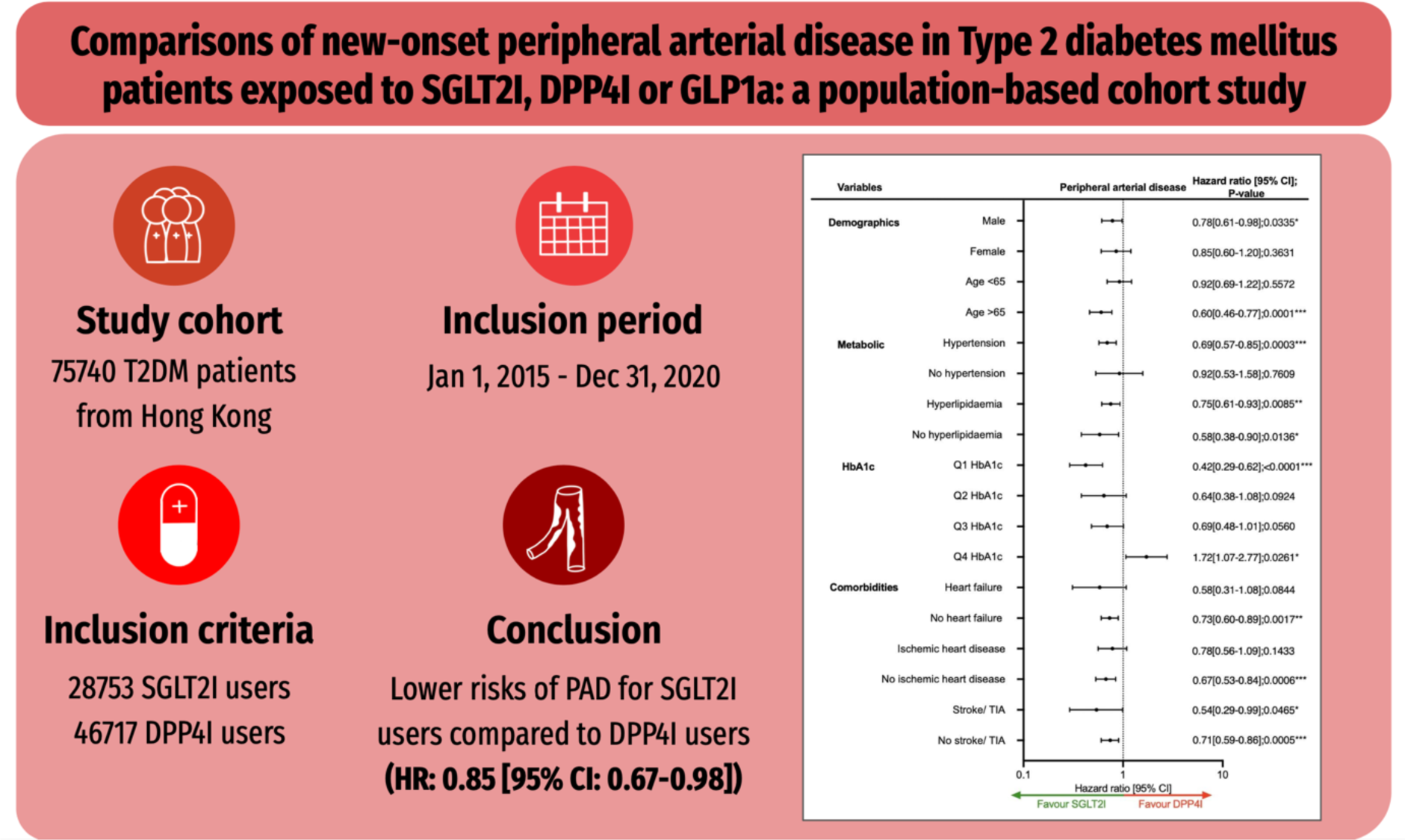

## Introduction

The rising incidence of type 2 diabetes mellitus (T2DM) is a major global concern, imposing a significant burden on economies and healthcare systems worldwide [1]. Among the most prevalent adverse events related to T2DM is peripheral arterial disease (PAD), culminating in ulcerations, limb gangrene, and amputation[2]. Compared to coronary artery disease and stroke [3, 4], PAD studies are often underrepresented. Despite relatively lower incidence rates amongst Asian compared to European, a rising trend in the prevalence of PAD outcomes has been observed [5].

The use of anti-diabetic medications in treating peripheral artery disease (PAD) has garnered significant interest. Several studies have assessed the relationship between anti-diabetic drugs and atherosclerosis, demonstrating the propensity of certain classes to suppress such pathogenesis through their anti-inflammatory effects [6]. This further propagated interest on novel anti-diabetic medications such as sodium-glucose cotransporter 2 inhibitors (SGLT2I), dipeptidyl peptidase 4 inhibitors (DPP4I) and glucagon-like peptide-1 receptor agonist (GLP1a) on PAD outcomes, as these drugs are commonly prescribed as second-line anti-diabetic medications. Recent comparative studies have demonstrated the superiority of SGLT2I relative to DPP4I in reducing the risk of adverse lower limb events in patients with existing PAD [7].

It is important to determine whether anti-diabetic drugs can lower the risk of PAD outcomes. This may encourage clinicians to reevaluate treatment for patients at risk of PAD outcomes. Previously it was suggested that SGLT2I was associated with a higher risk of PAD compared to GLP1a, while no statistical differences when compared to DPP4I [8]. Currently, clinical data surrounding the association of SGLT2I and DPP4I on PAD outcomes remain scarce. Hence, this study aimed to investigate the role of SGLT2I, DPP4I, and GLP1a with new-onset PAD-related outcomes in a cohort of T2DM patients from a territory-wide population cohort study.

## Methods

### Study population

This study was approved by the Institutional Review Board of the University of Hong Kong/Hospital Authority Hong Kong West Cluster (HKU/HA HKWC IRB) (UW-20-250) Clinical Research Ethics Committee and complied with the Declaration of Helsinki. This was a retrospective population-based study of prospectively collected electronic health records using the Clinical Data Analysis and Reporting System (CDARS) by the Hospital Authority (HA) of Hong Kong. The records cover both public hospitals and their associated outpatient clinics as well as ambulatory and daycare facilities in Hong Kong. This system has been used extensively by our teams and other research teams in Hong Kong, including diabetes and medication research [9, 10]. The system contains data on disease diagnosis, laboratory results, past comorbidities, clinical characteristics, and medication prescriptions. T2DM patients who were administered with SGLT2I or DPP4I in centres under the HA, between 1^st^ January 2015, to 31^st^ December 2020, were included. The GLP1a cohort comprised of patients on GLP1a between 1^st^ January 2015, to 31^st^ December 2020 was included for sensitivity analysis to demonstrate the relative effects amongst the second-line oral anti-diabetic agents.

### Predictors and variables

Patients’ demographics include gender and age of initial drug use (baseline), clinical and biochemical data were extracted for the present study. Prior comorbidities were extracted by the *International Classification of Diseases Ninth Edition* (ICD-9) codes (**Supplementary Table 1**). The diabetes duration was calculated by examining the earliest date amongst the first date of (1) diagnosis using ICD-9; (2) HbA1c ≥6.5%; (3) Fasting glucose ≥7.0 mmol/l or Random glucose 11.1 mmol/l; (4) using anti-diabetic medications. The patients on financial aid were defined as patients on Comprehensive Social Security Assistance (CSSA) scheme, higher disability allowance, normal disability allowance, wavier, other financial aids in Hong Kong. The number of hospitalisations in the year prior to the index days was extracted. The Charlson standard comorbidity index was calculated [11]. The duration and frequency of SGLT2I and DPP4I usage was calculated. The baseline laboratory examinations, including the glucose profiles and renal function tests, were extracted. The estimated glomerular filtration rate (eGFR) was calculated using the abbreviated modification of diet in renal disease (MDRD) formula [12]. Furthermore, the time-weighted lipid and glucose profiles after drug initiation were also calculated by the products of the sums of two consecutive measurements and the time interval, then divided by the total time interval, as suggested previously [13].

The following patient groups were excluded: (1) those with prior PAD (2) patients who died within 30 days after initial drug exposure; (3) without complete demographics; (4) those under 18 years old; (5) developed PAD within 30 days after drug exposure (**Figure 1**). To account for the incomplete laboratory data, the multiple imputation by chained equations was performed according to a previously published study [14].

**Figure 1.**
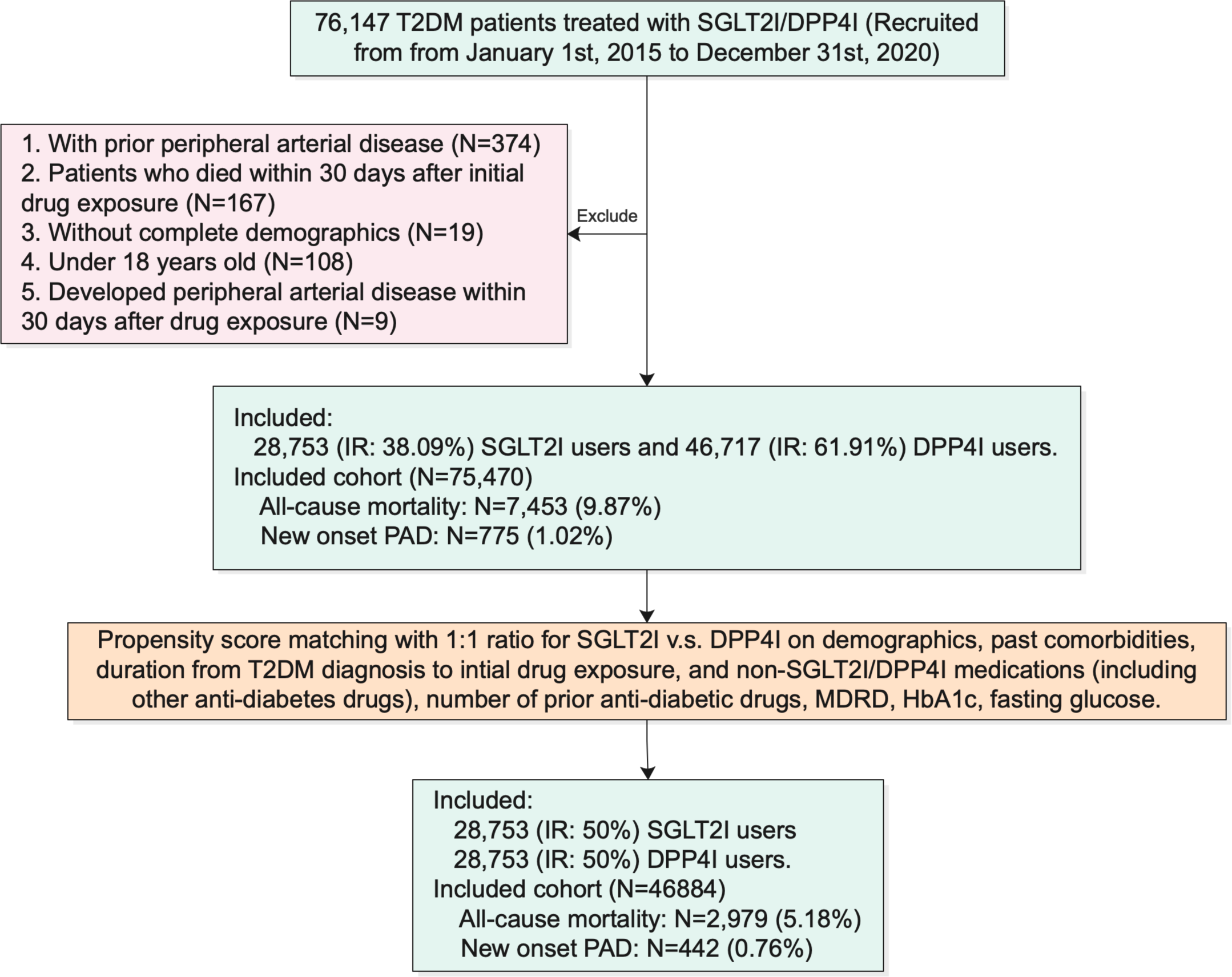
Procedures of data processing for the study cohort. SGLT2I: Sodium-glucose cotransporter-2 inhibitors; DPP4I: Dipeptidyl peptidase-4 inhibitors. MDRD: modification of diet in renal disease. PAD: Peripheral Arterial Disease

### Study outcomes

The primary outcome of this study was new-onset PAD defined using either ICD-9 code, history of amputation due to PAD, and peripheral arterial intervention (**Supplementary Table 1**), according to the verified published literature [15]. The secondary outcome was all-cause mortality. Mortality data were obtained from the Hong Kong Death Registry, a population-based official government registry with the registered death records of all Hong Kong citizens linked to CDARS. Mortality was recorded using the *International Classification of Diseases Tenth Edition* (ICD-10) coding. The as-treat approach was adopted which patients were censored at treatment discontinuation or switching of the comparison medications. The endpoint date of interest for eligible patients was the event presentation date. The endpoint for those without primary outcome was the mortality date or the end of the study period (31^st^ December 2020).

### Statistical analysis

Descriptive statistics are used to summarize baseline clinical and biochemical characteristics of patients with SGLT2I and DPP4I use. For baseline clinical characteristics, continuous variables were presented as mean (95% confidence interval/standard deviation), and the categorical variables were presented as total numbers (percentage). Propensity score matching generated by logistic regression with 1:1 ratio for SGLT2I use versus DPP4I use based on demographics, non-SGLT2I/DPP4I medications, number of prior anti-diabetic drugs, prior comorbidities, renal function, duration from T2DM diagnosis initial drug exposure, HbA1c and fasting glucose were performed using the nearest neighbour search strategy with a calliper of 0.1. Propensity score matching was performed using Stata software (Version 16.0). Baseline characteristics between patients with SGLT2I and DPP4I use before and after matching were compared with absolute standardized mean difference (SMD), with SMD<0.10 regarded as well-balanced between the two groups.

The cumulative incidence curves for the primary outcomes and secondary outcomes were constructed. Proportional Cox regression models were used to identify significant risk predictors of adverse study outcomes in the matched cohort, with adjustments for demographics, comorbidities, number of prior hospitalisations, medication profile, renal function, glycaemic tests, and the duration of T2DM. The log-log plot was used to verify the proportionality assumption for the proportional Cox regression models. Subgroup analyses were conducted to confirm the association amongst patients with different clinically important predictors accounting to the diabetic and the metabolic profile, as well as the comorbidities and medications associated with PAD.

Cause-specific and sub-distribution hazard models were conducted to consider possible competing risks. Multiple propensity adjustment approaches were used, including propensity score stratification[16], propensity score matching with inverse probability of treatment weighting (IPTW) [17] and propensity score matching with stable inverse probability weighting [18]. The three arm sensitivity results involving glucagon-like peptide-1 receptor agonist (GLP1a) using stabilized IPTW were conducted to test the association and choice amongst the novel second-line anti-diabetic medications. Sensitivity analyses result with consideration of one-year lag time effects was conducted. Patients with chronic kidney disease (CKD) stage 4/5 (eGFR <30 mL/min/1.73m*2), peritoneal dialysis or haemodialysis who may be contraindicated with SGLT2I were excluded from the sensitivity analysis. The hazard ratio (HR), 95% CI, and P-value were reported. Statistical significance was defined as P-value <0.05. All statistical analyses were performed with RStudio (Version: 1.1.456) and Python (Version: 3.6).

## Results

In this territory-wide cohort study of 75470 patients with T2DM treated with SGLT2I/DPP4I between 1^st^ January 2015 and 31^st^ December 2020 in Hong Kong, patients were followed up until 31^st^ December 2020 or until their deaths (**Figure 1**). The following patient groups were excluded: (1) prior PAD (N=334); (2) died within 30 days after initial f drug exposure (N=167); (3) without complete demographics (N=19); (4) under 18 years old (N=108); (5) developed PAD within 30 days after drug exposure (N=9).

After exclusions, this study included a total of 75470 patients with T2DM (mean age: 62.3 years old [SD: 12.8]; 55.79% males), of whom 28753 patients (38.09%) used SGLT2Is, and 46717 patients (61.97%) used DPP4Is (**Table 1**). Before matching, the SGLT2 users were comprised of more male patients, younger, with less comorbidities, with more cardiovascular diseases (hypertension and ischaemic heart disease), using more anti-diabetic drugs, had a higher eGFR compared to DPP4I users, lower high-density lipoprotein, and higher mean HbA1C and fasting glucose.

**Table 1.**
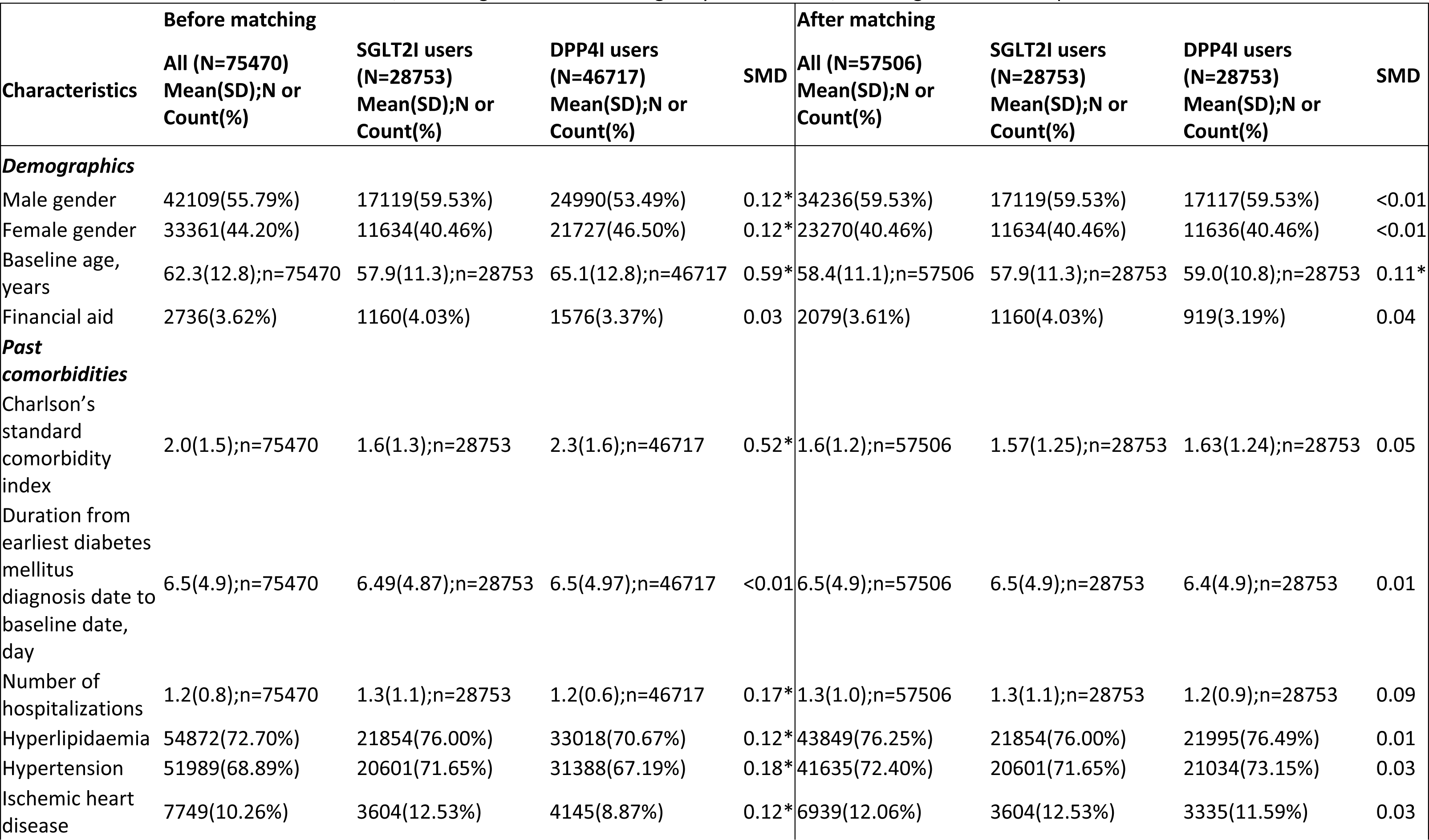
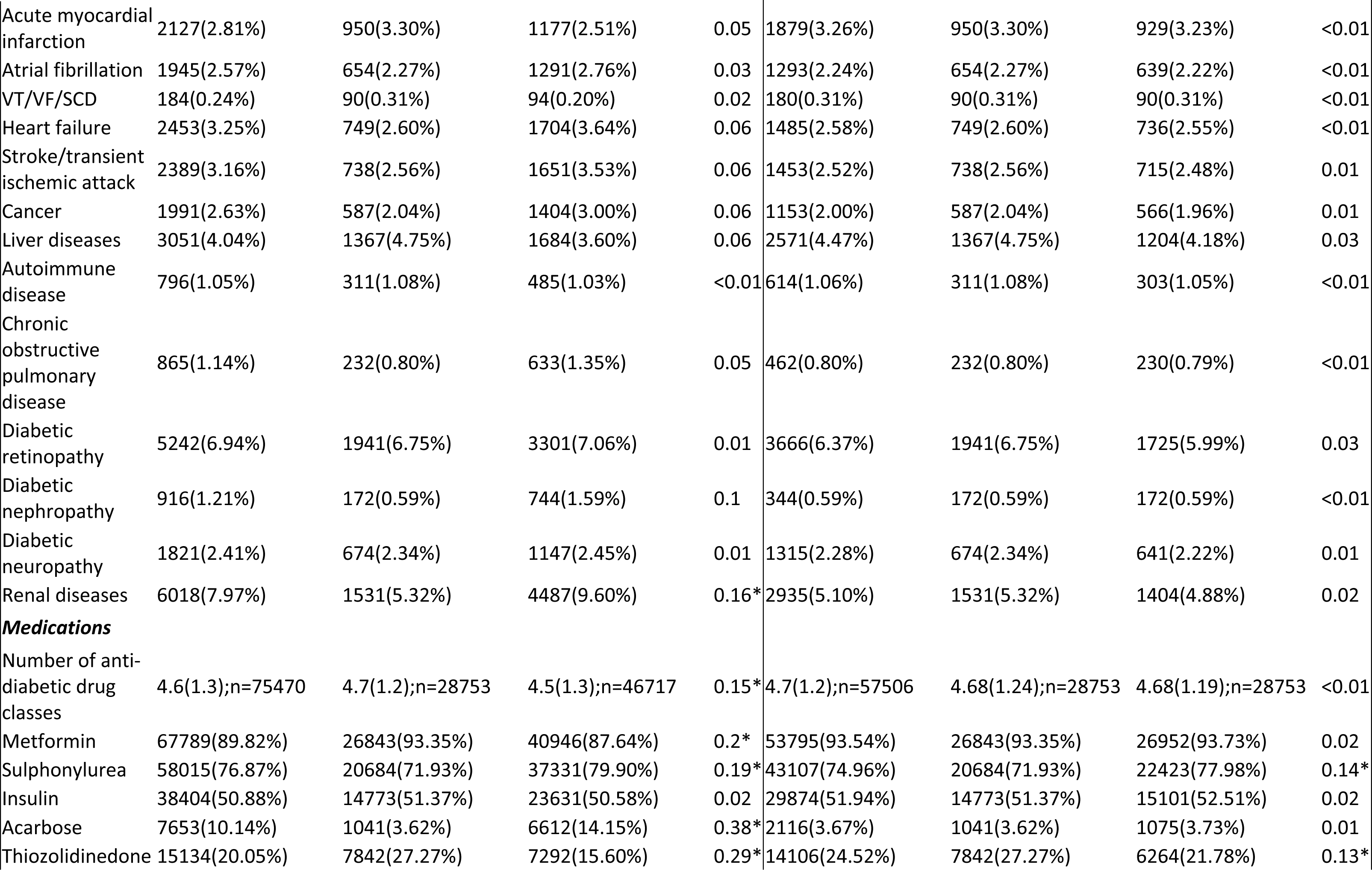
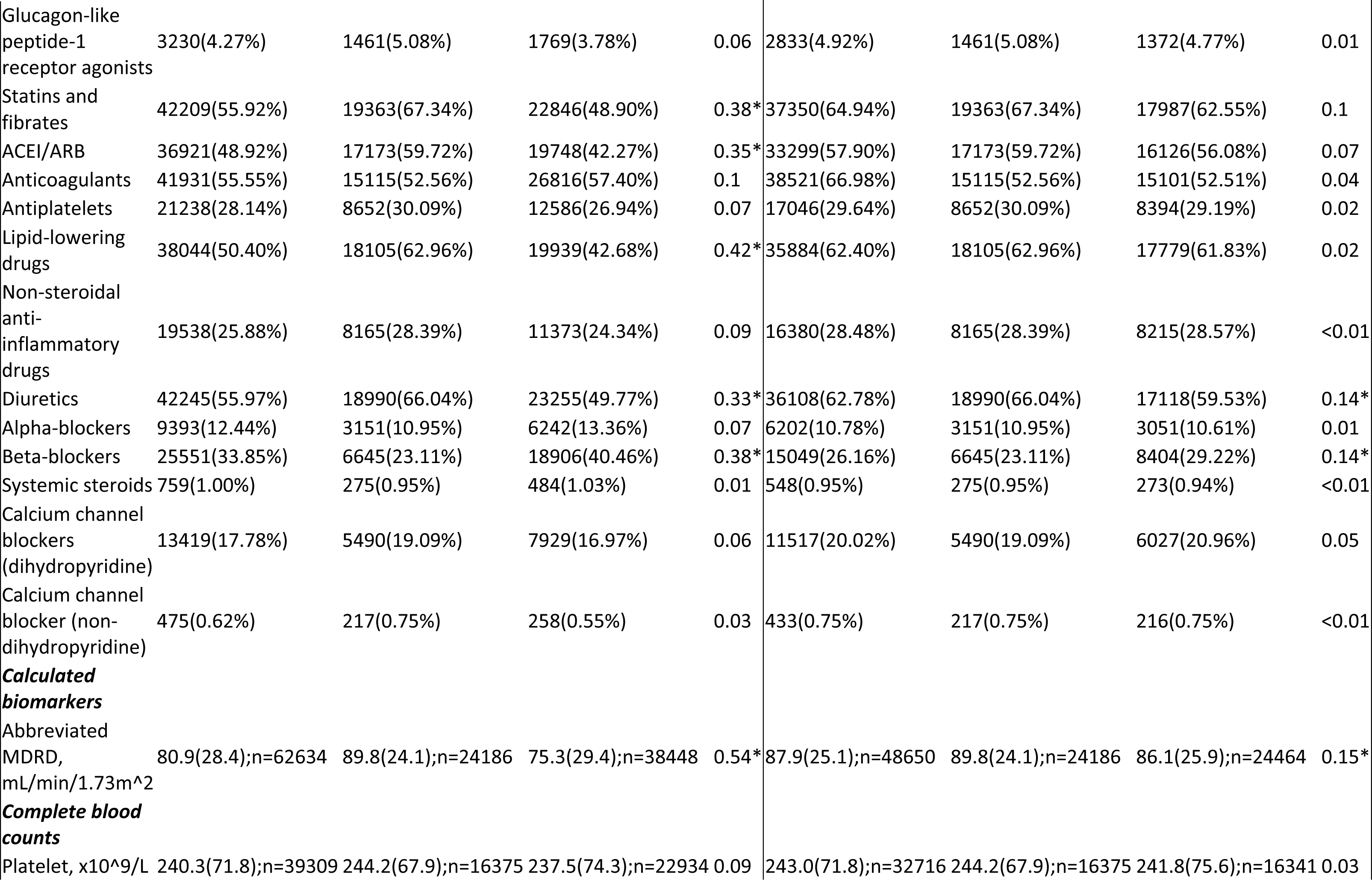
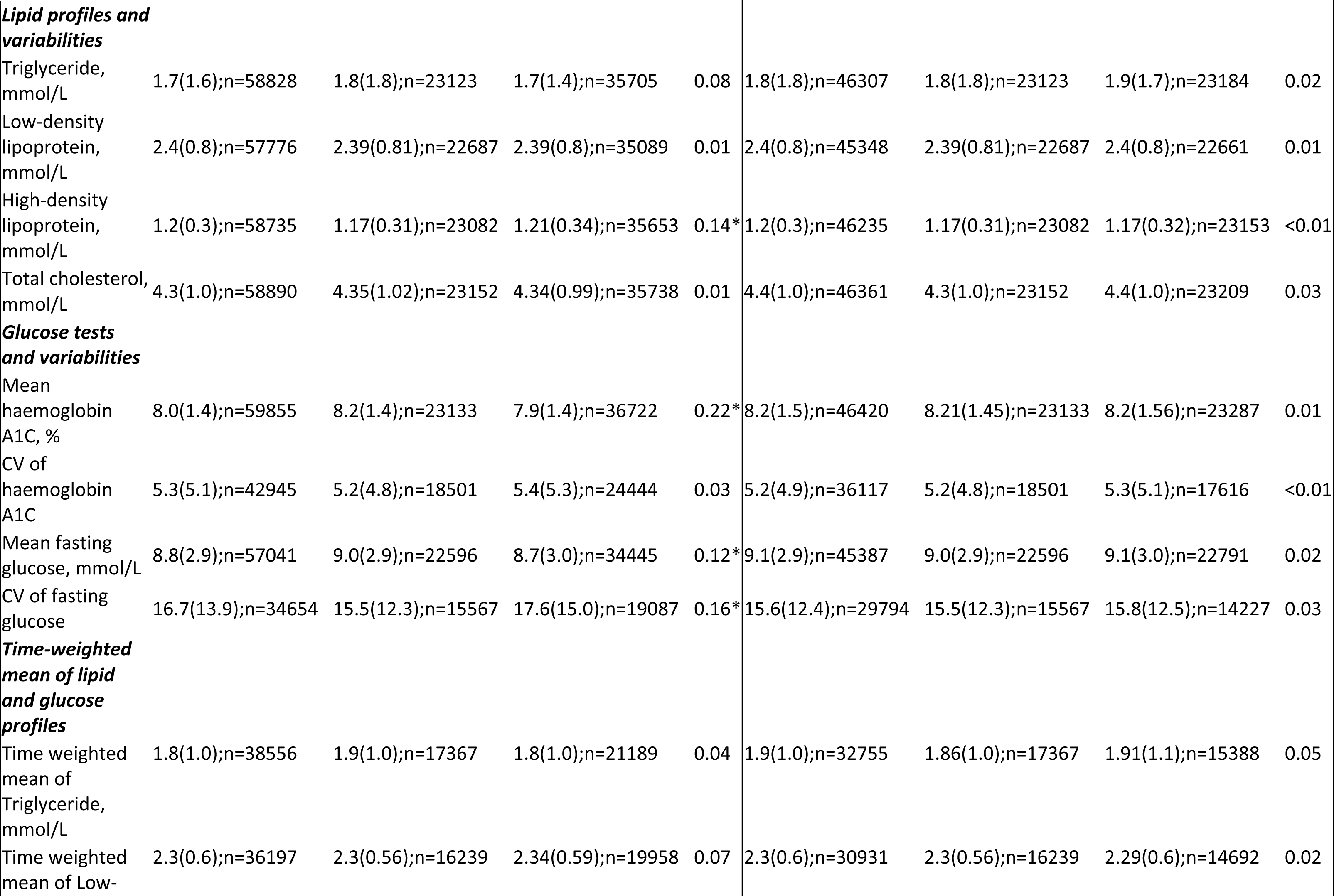
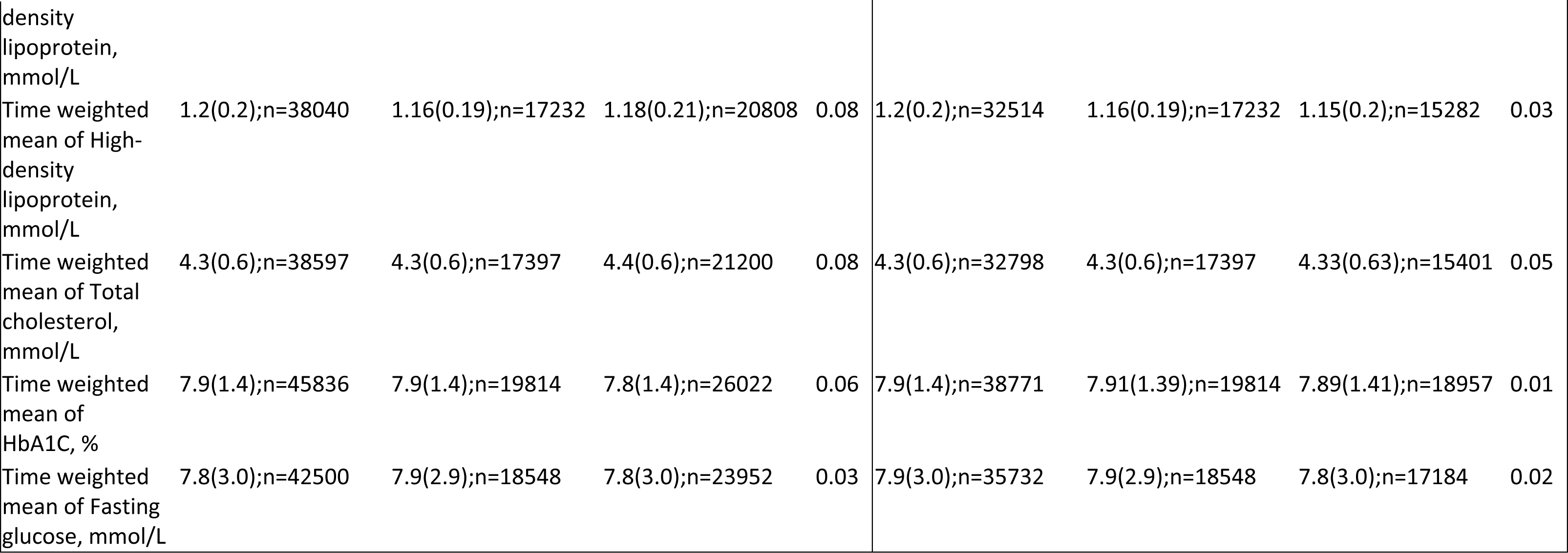
Baseline and clinical characteristics of patients with SGLT2I v.s. DPP4I use before and after propensity score matching (1:1). * for SMD≥0.1; SGLT2I: sodium glucose cotransporter-2 inhibitor; DPP4I: dipeptidyl peptidase-4 inhibitor; SD: standard deviation; CV: Coefficient of variation; MDRD: modification of diet in renal disease; ACEI: angiotensin-converting enzyme inhibitors; ARB: angiotensin II receptor blockers.

After the propensity score matching, baseline characteristics and the time-weighted lipid and glucose profiles of the 2 groups were well-balanced, apart from baseline age (SMD=0.11), sulphonylurea (SMD=0.14), thiazolidinedione (SMD=0.13), diuretics (SMD=0.14), beta-blocker (SMD=0.14), abbreviated MDRD (SMD=0.15) (**Table 1**). The DPP4I and SGLT2I cohorts were comparable after matching with nearest neighbour search strategy with calliper of 0.1, and the proportional hazard assumption was confirmed **(Supplementary** Figure 1). In the matched cohort, 442 patients developed PAD; 2979 patients passed away during the study period (**Figure 1**). The characteristics of patients are shown in **Table 1**.

### Association between SGLT2I and DPP4I and peripheral arterial diseases

In the matched cohort, 186 SGLT2I users and 256 DPP4I users developed PAD. After a mean follow-up of 314442.4 person-year, the incidence of PAD was lower amongst SGLT2I users (Incidence rate [IR] per 1000 person-year: 1.17; 95% CI: 1.01-1.35) compared to DPP4I users (IR per 1000 person-year: 1.65; 95% CI: 1.45-1.86) (**Table 2**). SGLT2I users had a 15% lower risk of PAD after adjustment (HR: 0.85; 95% CI: 0.67-0.98, p=0.0464) compared to users of DPP4I regardless of the demographics, comorbidities, medication profile, renal function, glycaemic tests, number of hospitalisations, and the duration of T2DM (**Table 2; Supplementary** Figure 2**; Supplementary Table 3)**. This was substantiated by the cumulative incidence curves stratified by SGLT2I versus DPP4I (**Figure 2**).

**Table 2.** Incidence rate (IR) per 1000 person-year and multivariate Cox regression models of new onset peripheral arterial disease (PAD), and all-cause mortality in the cohort before and after 1:1 propensity score matching. * for p≤ 0.05, ** for p ≤ 0.01, *** for p ≤ 0.001; CI: confidence interval; SGLT2I: sodium glucose cotransporter-2 inhibitor; DPP4I: dipeptidyl peptidase-4 inhibitor. Adjusted for demographics, past comorbidities, duration of diabetes mellitus, and number of hospitalizations, number of anti-diabetic drugs, non-SGLT2I/DPP4I medications, abbreviated MDRD, HbA1c, fasting glucose.

| Peripheral vascular diseases | Number of patients | Number of events | Total person-year | Follow-up duration, years [95% CI] | Incidence [95% CI] | Adjusted hazard ratio [95% CI] | P-value |
| --- | --- | --- | --- | --- | --- | --- | --- |
| **DPP4I** | 28753 | 256 | 155511.5 | 5.58(5.28-5.81) | 1.65[1.45-1.86] | 1 [Reference] | NA |
| **SGLT2I** | 28753 | 186 | 158930.9 | 5.62(5.35-5.82) | 1.17[1.01-1.35] | 0.85[0.67-0.98] | 0.0464* |
| All-cause mortality | Number of patients | Number of events | Total person-year | Follow-up duration, years [95% CI] | Incidence [95% CI] | Adjusted hazard ratio [95% CI] | P value |
| **DPP4I** | 28753 | 2129 | 156232 | 5.59(5.29-5.81) | 13.63[13.05-14.22] | 1 [Reference] | NA |
| **SGLT2I** | 28753 | 850 | 159497.5 | 5.62(5.35-5.83) | 5.33[4.98-5.70] | 0.59[0.54-0.64] | <0.0001*** |

**Figure 2.**
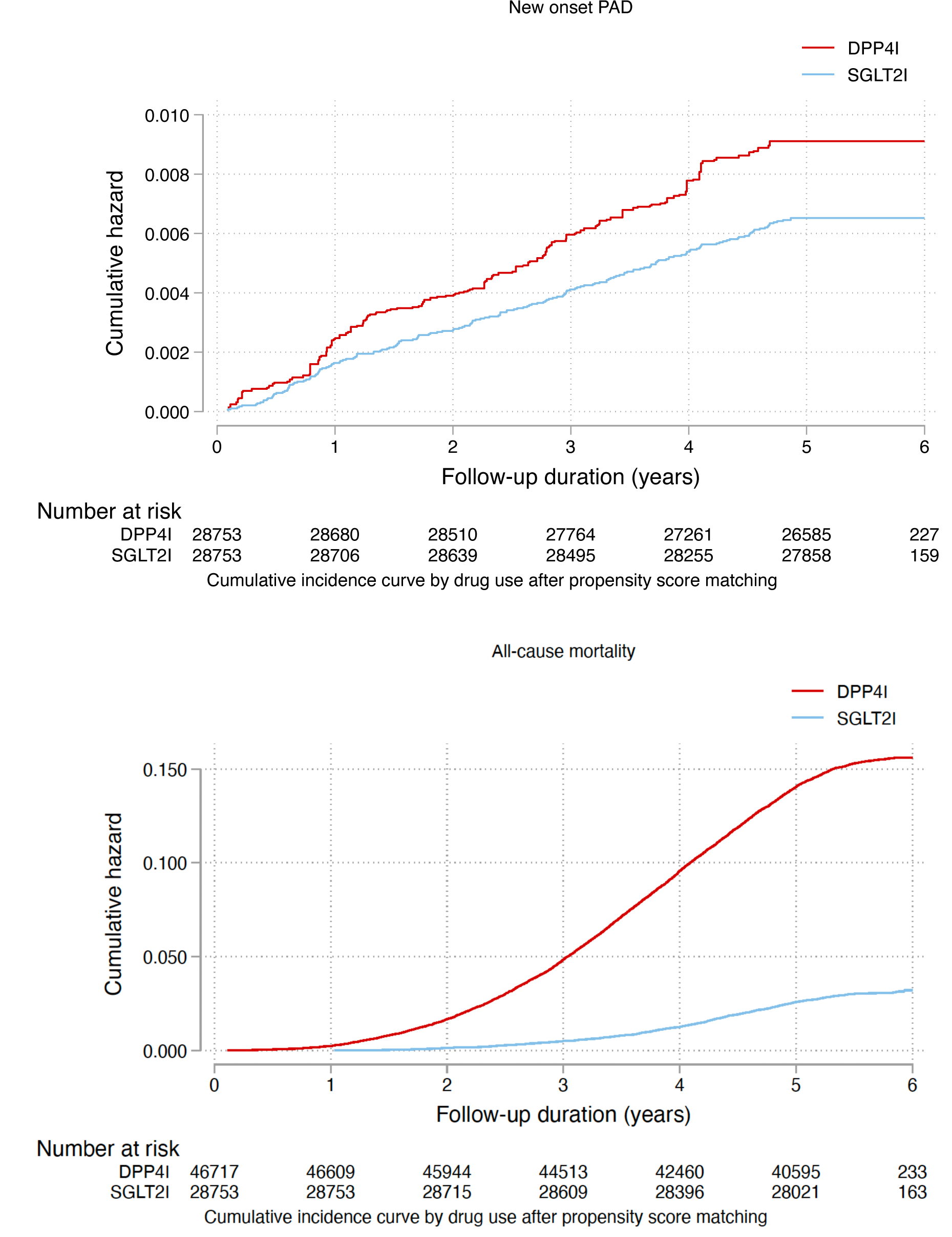
Cumulative incidence curves for new onset Peripheral Arterial Disease (PAD), and all-cause mortality stratified by drug exposure effects of SGLT2I and DPP4I after propensity score matching (1:1). SGLT2I: Sodium-glucose cotransporter-2 inhibitors; DPP4I: Dipeptidyl peptidase-4 inhibitors; PAD: Peripheral Arterial Disease

### Association between SGLT2I and DPP4I and all-cause mortality

Overall, 850 SGLT2I users and 2129 DPP4I users passed away. After a follow-up of 315729.5 person-year, the incidence of all-cause mortality was lower amongst SGLT2I users (IR: 5.33; 95% CI: 4.98-5.70) compared to DPP4I users (IR: 13.63; 95% CI: 13.05-14.22) (**Table 2**). SGLT2I users had a 41% lower risk of all-cause mortality after adjustment (HR: 0.59; 95% CI: 0.54-0.64, p<0.0001) compared to users of DPP4I regardless of the duration of diabetes mellitus (**Supplementary Table 3)**. This was substantiated by the cumulative incidence curves stratified by SGLT2I versus DPP4I (**Figure 2; Supplementary** Figure 2).

### Subgroup analysis

The results of the subgroup analysis for effects of SGLT2I and DPP4I on the PAD are shown in **Figure 3**. The result demonstrated that SGLT2I was associated with lower risks of PAD regardless of hyperlipidaemia, stroke/transient ischaemic attack, cancer, uses of ACEI/ARB, insulin, and metformin. SGLT2I was associated with lower risks of peripheral arterial disease only amongst male patients, patient older than 65 years old, with hypertension, without heart failure, without ischaemic heart diseases. Besides, SGLT2I was only associated with lower risks of peripheral arterial disease amongst patients currently on diuretics, lipid lowering drugs, anti-platelets, and anticoagulants, but not sulphonylurea, thiazolidinedione, and GLP1a. Regarding the diabetic control, SGLT2I was associated with lower risks of peripheral arterial disease only amongst the 1^st^ quartile of mean HbA1c. Interestingly, SGLT2I was associated with higher risks compared to DPP4I amongst those in the 4^th^ quartile of mean HbA1c. The marginal effects analysis demonstrated that SGLT2I was associated with lower risks of PAD consistently across all eGFR, number of anti-diabetic drugs used, and duration of diabetes, and the effects are more pronounced amongst patients with lower eGFR (**Supplementary** Figure 3).

**Figure 3.**
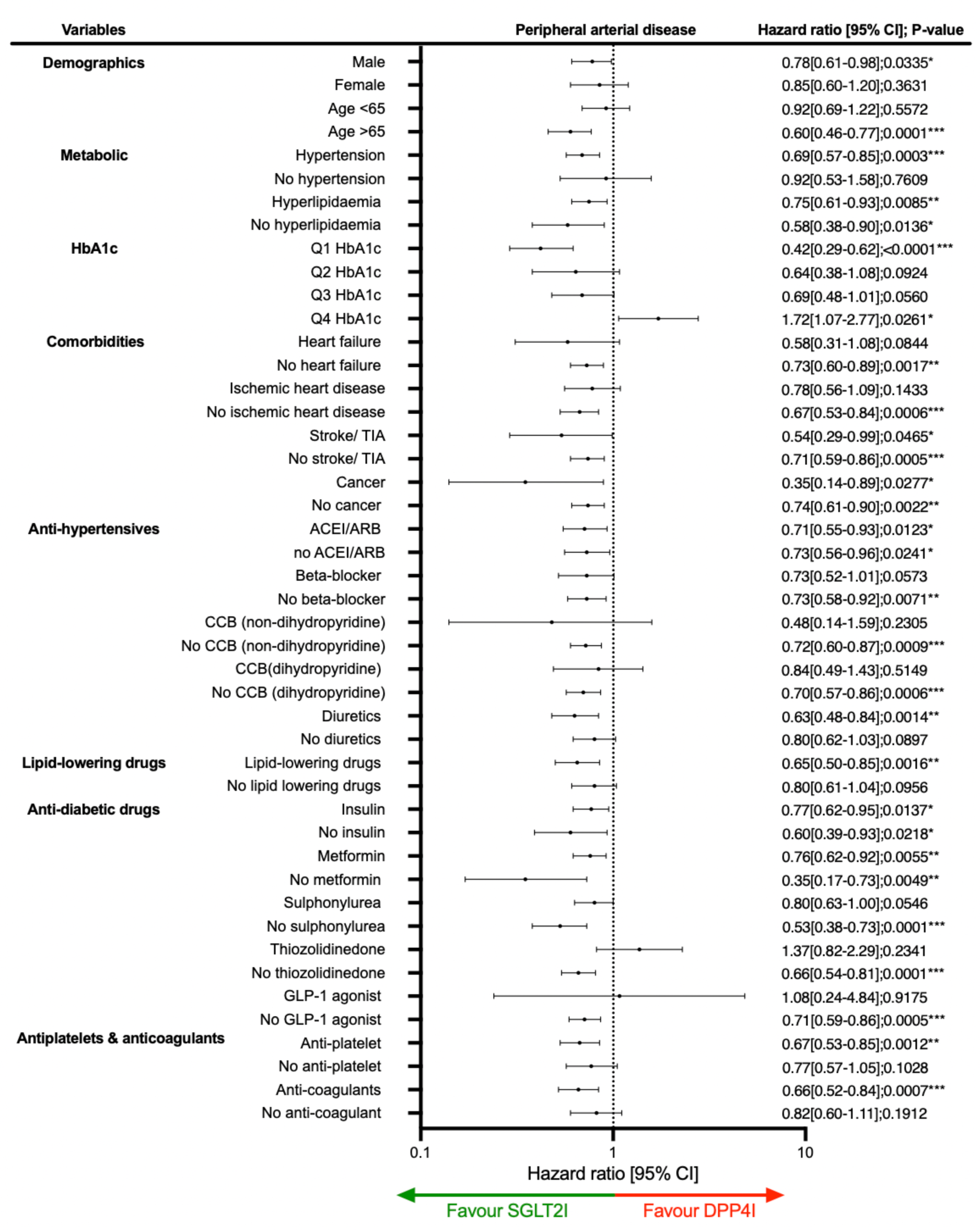
Forests plot of hazard ratios with 95% CI for SGLT2I v.s. DPP4I on new onset Peripheral Arterial Disease (PAD), and all-cause mortality stratified by drug use in the matched cohort. ACEI: angiotensin-converting enzyme inhibitors; ARB: angiotensin II receptor blockers; CCB: Calcium channel blocker; SGLT2I: Sodium-glucose cotransporter-2 inhibitors; DPP4I: Dipeptidyl peptidase-4 inhibitors; GLP1a: glucagon-like peptide-1 receptor agonist; TIA: Transient ischaemic attack

### Sensitivity analysis

Sensitivity analyses were performed to confirm the predictability of the models. The results of the cause-specific hazard models, sub-distribution hazard models and different propensity score approaches demonstrated that different models did not change the point estimates for both the primary and the secondary outcomes (all p<0.05) **(Supplementary Table 4)**. A 3-arm analysis with the inclusion of GLP1a including patients only on (SGLT2I, DPP4I, and GLP1a) using stabilized IPTW was conducted **(Supplementary Table 5)**. The result between DPP4I and SGLT2I remained consistent with the main result (HR: 0.72; 95% CI: 0.59-0.87). Meanwhile, GLP1a was not associated with higher risks of peripheral arterial disease (p>0.05). Excluding patients with CKD stage 4/5 (eGFR <30), peritoneal dialysis or haemodialysis in the matched cohort likewise demonstrated that SGLT2I was associated with lower risks of peripheral arterial disease outcomes compared to DPP4I **(Supplementary Table 6)**. The sensitivity analysis for 1-year lag time also demonstrated the same trend **(Supplementary Table 7)**. Lastly, a sensitivity analysis was conducted to exclude patients who were on financial aids, and the association between SGLT2I and PAD remained consistent **(Supplementary Table 8)**.

## Discussion

In this territory-wide cohort study, the relative associations between SGLT2I, DPP4I and new-onset PAD outcomes was examined using real-world data and offers novel insights in diabetic management. Our results firstly demonstrate that SGLT2I use was independently associated with a lowered risk of new-onset PAD than DPP4I usage after adjustments. Similar associations were observed in subgroup analyses in male patients above 65 years old, with hypertension, and low HbA1c levels. Secondly, SGLT2I was not associated with lower risks of PAD compared to GLP1a.

### Comparison with previous studies

The prevalence of PAD outcomes in our study for SGLT2I and for DPP4I parallels the values of existing studies, which described an incidence of 2.7 per 1000 person-year [19], lower than the Western counterparts in the ADVANCE trial (12 per 1000 person-year) [20]. In our study, SGLT2I users had relatively reduced risks of PAD outcomes compared to DPP4I users. It should be noted that the observed effect of SGLT2I in our cohort was marginal and may produce a minor modulating effect against new-onset PAD in diabetic patients.

The relationship between SGLT2I on coronary artery disease and cerebral vascular diseases has been well established in large randomised controlled trials recent years [21, 22]. However, the clinical data regarding the relationship between SGLT2I on PAD remain scarce. In our study, we show novel findings that SGLT2I was associated with lower risks of PAD compared to DPP4I.

In the subgroup analysis, the results demonstrated that in terms of PAD, SGLT2I was better amongst patients with low mean HbA1c level, but DPP4I was better amongst patients with high HbA1c levels. The difference extent of glycaemic control contributes to the systemic and renal arterial stiffness, and potentially explaining the differences in the association in PAD [23]. The use of SGLT2I also correlates with a reduction in blood pressure and CRP levels amongst diabetic patients [24]. A study applying the Archimedes model deduced that dapagliflozin usage reduced foot amputations by 13% relative to standard care [25].

Lin *et al.* demonstrated that canagliflozin correlated with increase in amputation risk and PAD incidence [26], and in the Canagliflozin Cardiovascular Assessment Study (CANVAS) trial the risk of amputation due to vascular disease even increased by twofold [27]. However, meta-analysis of the DAPA-HF and DELIVER trials suggested that dapagliflozin did not increase the risk of amputation compared to placebo regardless of PAD status [28]. These differences could be explained by the differences in the study population and design.

A Taiwanese cohort study found that GLP1a was associated with decreased risks of major adverse limb events and PAD compared to DPP4I [29]. In our sensitivity analysis, the risks of PAD were similar between SGLT2I and GLP1a **(Supplementary Table 5)**. Therefore, we hypothesized that GLP1a might also reduce the risks of PAD amongst T2DM patients, however the results were inconclusive provided the relatively small GLP1a sample size. Our results are aligned with existing analyses on SGTL2I and GLP1a on the risk of PAD amongst hospitalised patients with heart failure (HR: 0.89; 95% CI: 0.57-1.33) [30]. Also, a Danish cohort study showed that GLP1a users had a lower risk of lower limb amputation compared to sulfonylureas [31]. The relationship between SGLT2I and DPP4I with GLP1a all-cause mortality was generally consistent with the existing randomised controlled trials results [32].

However, the results do not necessarily mean that DPP4I increases the risks of new-onset PAD outcomes. It is still plausible to hypothesize that DPP4I may possess protective effects, although further studies are necessary to verify this proposition. DPP4I usage may lower the risk of limb amputation and PAD occurrence amongst diabetic patients [33], but there is limited evidence comparing the role of SGLT2I versus DPP4I on PAD and other crucial patient outcomes [7]. With the available evidence, the results are highly heterogenous and thus difficult to draw any definitive consensus.

On the other hand, a meta-analysis revealed little difference between SGLT2I, DPP4I and GLP1a or other types of oral hypoglycaemic agents on the prognosis of diabetic-foot related limb amputations [34]. One meta-analysis of 12 cohorts suggests that only GLP1a usage, but not SGLT2I, was associated with lower risk of lower limb amputation compared to DPP4I; however, no such correlation was identified with SGLT2I usage [35].

The mechanism(s) behind the relationship between SGLT2I and PAD remain unknown. It was suggested that canagliflozin was associated with enhanced vasodilation of coronary arteries in diabetic mice [36]. The existing literature attributes the possible pleiotropic effects of SGLT2I to the suppression of oxidative stress and autophagy regulation, which may minimize atherosclerosis [37]. SGLT2I usage might also activate NLRP3 inflammasome, associated with inflammatory signalling, thereby reducing the development of atherosclerosis [38]. Alternatively, some found that dapagliflozin could generate anti-inflammatory effects via stimulation of macrophages and cytokines [39].

### Clinical implications

The potential protective effects of SGLT2I and DPP4I on various forms of PAD outcomes have attracted increasing attention over recent years. As of 2017, international institutions such as the European Medicines Agency (EMA) issued official warnings against the use of the whole class of SGLT2I treatments due to the potential concern for elevated risks of lower extremity minor and major amputations [30]. Hence, our study results along with existing literature may inspire re-evaluation of current guidelines regulating the use of novel anti-diabetic medications for patients who are at higher risk of developing PAD outcomes. Using data from clinical practice in Hong Kong, our preliminary findings suggest that SGLT2I may exhibit a possible beneficial effect against PAD relative to DPP4I usage. Although it is difficult to establish any definitive claims at this stage, our study may prompt more random clinical trial studies investigate this association. In the long term, this may produce a paradigm shift in diabetic management which have traditionally cantered around glycaemic regulation, to assess the potential secondary benefits of novel anti-diabetic medications for patients at risk of PAD.

### Limitations

Several limitations present in this study merits consideration. The observational nature of this study suggests that data on certain variables namely alcohol use, BMI, smoking, family history of PAD and physical activity could not be obtained and analyzed. In addition, the data may also be susceptible to coding errors, under-coding and missing data, resulting in information bias. In compensation of this, laboratory results and comorbidities related to PAD were used to infer for possible risk factors. To corroborate, drug cohorts were matched over a variety of medications and diseases, adjusted with regression, and performed sensitivity analyses to reduce the effect of bias. Moreover, one concern in data on PAD was that the condition was often underdiagnosed as most patients may not present to the hospital until heavily symptomatic. In acknowledgement of this, our study identified clinically significant PAD patients that required medical attentions, amputation, or revascularization. Furthermore, the retrospective nature of the study design suggests that all results derived from our study regarding the relationship between SGLT2I, DPP4I and PAD remain correlational. Hence, further randomized clinical trials are needed to substantiate the potential causal relationship and the consequences of undiagnosed PAD patients.

## Conclusion

This population-based cohort study suggested that SGLT2I users were associated with lower risks of new-onset PAD and PAD-related outcomes amongst T2DM patients. Further investigations are warranted to verify the effects of SGLT2I or other anti-diabetic medications on PAD.

## Data Availability

An anonymised version without identifiable or personal information is available from the corresponding authors upon reasonable request for research purposes.

## Acknowledgements

None.

## Ethical approval statement

This study was approved by the Institutional Review Board of the University of Hong Kong/Hospital Authority Hong Kong West Cluster (HKU/HA HKWC IRB) (UW-20-250), and New Territories East Cluster-Chinese University of Hong Kong (NTEC-UCHK) Clinical Research Ethnics Committee (2018.309, 2018.643) and complied with the Declaration of Helsinki.

## Guarantor Statement

All authors approved the final version of the manuscript. GT is the guarantor of this work and, as such, had full access to all the data in the study and takes responsibility for the integrity of the data and the accuracy of the data analysis.

## Author contributions

Data analysis: OHIC, ZL, HL, JZ

Data review: OHIC, ZL, SL, GT, JZ

Data acquisition: OHIC, SL

Data interpretation: OHIC, ZL, CTC, JC, HL, IL, JZ

Critical revision of manuscription: QZ, TL, BMYC, WTW, GL, IW, GT, JZ

Supervision: BMYC, GT, FPL, JZ

Manuscript writing: OHIC, CTC, JC, IL

Manuscript revision: OHIC, ZL, CTC, JC, GL, IW, GT, FPL, JZ

