## Supplementary Appendix for "Comparisons of new-onset peripheral arterial disease in Type 2 diabetes mellitus patients exposed to SGLT2I, DPP4I or GLP1a: a population-based cohort study"

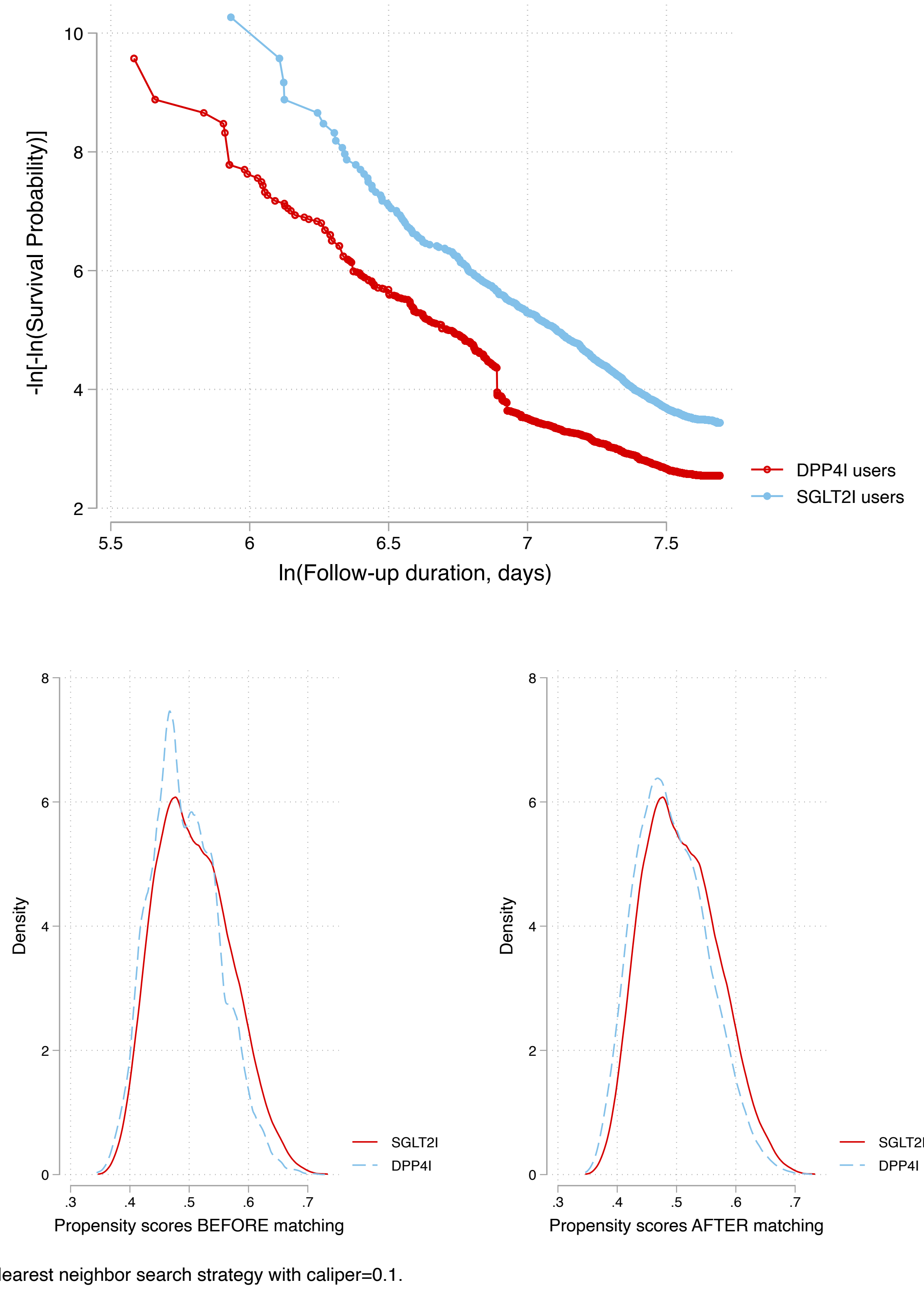

**Supplementary Figure 1. Propensity score matching comparisons and proportional hazard assumption checking with parallel lines for SGLT2I versus DPP4I before and after 1:1 matching with nearest neighbour search strategy with calliper of 0.1**

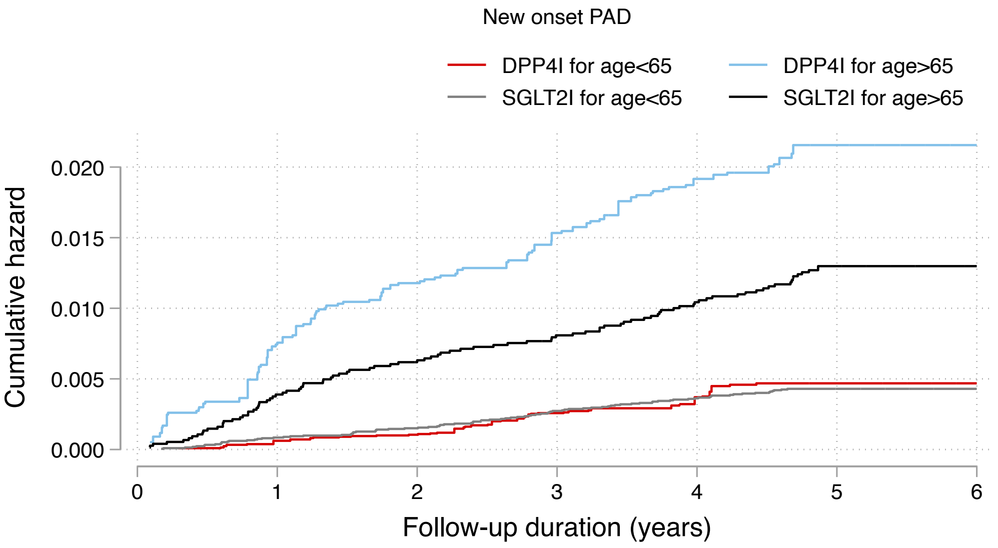

Number at risk

| | 0 | 1 | 2 | 3 | 4 | 5 | 6 |
|---|---|---|---|---|---|---|---|
| DPP4I for age<65 | 21059 | 21044 | 21013 | 20564 | 20378 | 20135 | 176 |
| DPP4I for age>65 | 7694 | 7636 | 7497 | 7200 | 6883 | 6450 | 51 |
| SGLT2I for age<65 | 21288 | 21270 | 21246 | 21179 | 21081 | 20914 | 115 |
| SGLT2I for age>65 | 7465 | 7436 | 7393 | 7316 | 7174 | 6944 | 44 |

Cumulative incidence curve by drug use after propensity score matching

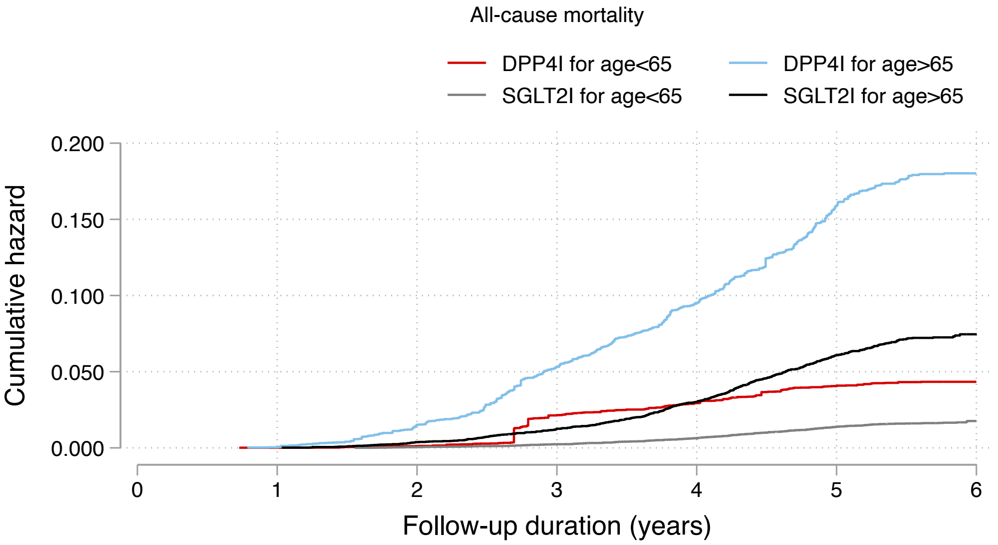

Number at risk

| | 0 | 1 | 2 | 3 | 4 | 5 | 6 |
|---|---|---|---|---|---|---|---|
| DPP4I for age<65 | 21059 | 21057 | 21034 | 20612 | 20448 | 20220 | 176 |
| DPP4I for age>65 | 7694 | 7691 | 7578 | 7296 | 6996 | 6564 | 52 |
| SGLT2I for age<65 | 21288 | 21288 | 21278 | 21237 | 21153 | 20996 | 116 |
| SGLT2I for age>65 | 7465 | 7465 | 7437 | 7372 | 7243 | 7025 | 47 |

Cumulative incidence curve by drug use after propensity score matching

**Supplementary Figure 2. Cumulative incidence curves for new onset PAD, and all-cause mortality stratified by patient age at initial drug exposure and the combinations of age and drug exposure effects of SGLT2I and DPP4I in the matched cohort (Propensity score matching ratio 1:1)**

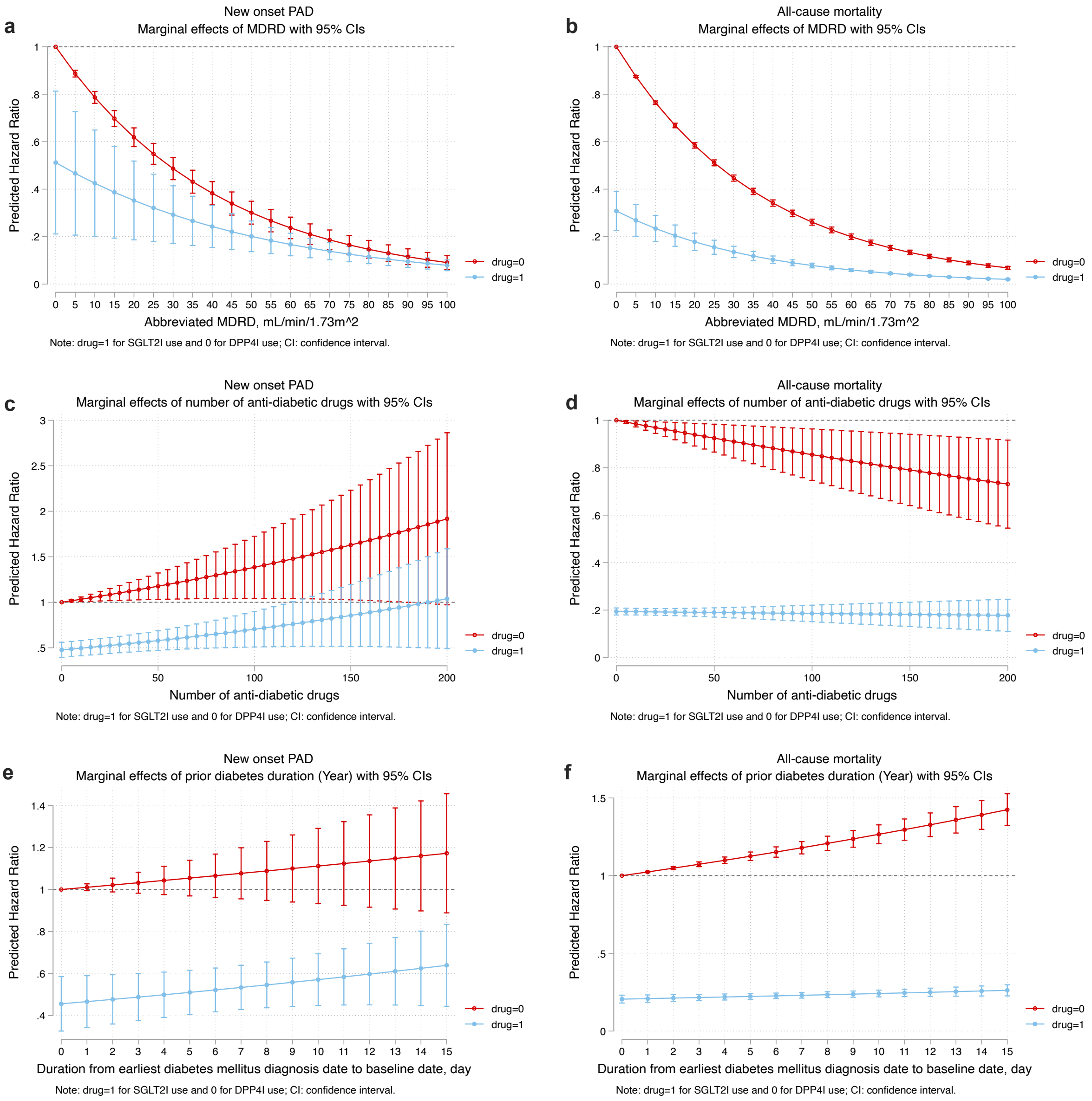

**Supplementary figure 3. Marginal effects of MDRD, number of prior anti-diabetic drugs, and prior diabetes duration with 95% Cis on new onset Peripheral Arterial Disease (PAD), and all-cause mortality stratified by drug use in the matched cohort.**

SGLT2I: Sodium-glucose cotransporter-2 inhibitors; DPP4I: Dipeptidyl peptidase-4 inhibitors.

**Supplementary Table 1. The International Classification of Diseases, Clinical Modification (ICD-9-CM) codes for definitions of past comorbidities and outcomes.**

| **Adverse outcome of interest** |
|---|
| Peripheral vascular disease: 249.70 249.71 250.70 250.71 250.72 250.73 440.2 440.21 440.22 440.23 440.24 440.29 440.3 440.30 440.31 440.32 440.8, 440.9, 443.81, 443.9, 444.22, 444.81<br>- Procedure: 00.55 00.60 39.90<br>- Amputation (1): 84.1 84.91 exclude concomitant 170.6-170.9, 171.3, 172.7, 173.7, 198.5, 344.1, 711.0, 728.86, 733.2, 736.3×-736.9, 735.×, 754.3×-754.7×,755.02, 755.13, 755.14, 755.3, 755.4, 755.6×, 755.8, 759.7, 759.89, 820.××-829.××, 835.××-838.××, 890.××, 891, 895.××-897.××, 904.××, 905.4, 928.××, 929.××,959.6, 959.7,996.4×, 996.66, 996.67, 996.77, 996.78 |
| **Past comorbidities** |
| **Cancer**: 140-239 |
| **Hypertension**: 401 401.1 401.9 402 402.01 402.1 402.11 402.9 402.91 403 403.01 403.1 403.11 403.9 403.91 404 404.01 404.02 404.03 404.1 404.11 404.12 404.13 404.9 404.91 404.92 404.93 405 405.01 405.09 405.1 405.11 405.19 405.9 405.91 405.99 437.2 + history of uses of anti-hypertensives |
| **Hyperlipidaemia**: 272.0 272.1 272.2 272.3 272.4 + history of uses of lipid-lowering drugs + lipid laboratory results |
| **Liver disease:** 275.1 275.0 572.0 572.4 572.1 572.3 572.8 573.0 573.4 573.8 573.9 |
| **Gastrointestinal bleeding:** 578.9 |
| **Heart failure:** 428 428 428.1 428.2 428.2 428.21 428.22 428.23 428.3 428.3 428.31 428.32 428.33 428.4 428.4 428.41 428.42 428.43 428.9 398.91 402.01 402.11 402.91 404.01 404.03 404.11 404.13 404.91 404.93 |
| **Atrial fibrillation:** 427.31 429.4 |
| **Stroke/transient ischemic attack:** 435 435.1 435.2 435.3 435.8 435.9 433.81 433.91 434 436 437 437.1 433.31 433.01 434.01 434.1 434.11 434.9 434.91 437.2 437.3 437.4 437.5 437.6 437.7 437.8 437.9 430 431 432 432.1 432.9 |
| **Ischemic heart disease:** 410.01 410.02 410.1 410.11 410.12 410.2 410.21 410.22 410.3 410.31 410.32 410.4 410.41 410.42 410.5 410.51 410.52 410.6 410.61 410.62 410.7 410.71 410.72 410.8 410.81 410.82 410.9 410.91 410.92 411 411.1 411.8 411.81 411.89 413 413.1 413.9 414 414.01 414.02 414.03 414.04 414.05 414.06 414.07 414.1 414.11 414.12 414.19 414.2 414.3 414.4 414.8 414.9 410 412 |
| **Acute myocardial infarction:** 410 410.01 410.02 410.1 410.11 410.12 410.2 410.21 410.22 410.3 410.31 410.32 410.4 410.41 410.42 410.5 410.51 410.52 410.6 410.61 410.62 410.7 410.71 410.72 410.8 410.81 410.82 410.9 410.91 410.92 |
| **Renal diseases:** 582 582 582.1 582.2 582.4 582.8 582.81 582.89 582.9 583 583 583.1 583.2 583.4 583.6 583.7 585 585.1 585.2 585.3 585.4 585.5 585.6 585.9 586 588 588 588.1 588.8 588.81 588.89 588.9 |
| **Diabetic retinopathy:** 250.5 361 362.01 362.02 362.1 362.53 362.81 362.82 362.83 369 379.23 |
| **Diabetic nephropathy:** 250.40 250.41 250.42 250.43 |
| **Diabetic neuropathy**: 250.6, 337.0, 337.1, 354.0 – 355.9, 356.9, 357.2, 358.1, 536.3, 564.5, 596.54, 713.5, 951.0, 951.1, 951.3 |
| **Chronic obstructive pulmonary disease** 491.0 491.1 491.2 491.8 491.9 492.0 492.8 496 |

**Supplementary Table 2. Calculations for variability measure**

| **Variability measure** | **Definition** |
|---|---|
| Coefficient of variation (CV) | $\frac{SD}{individual\ mean}$ |

.

**Supplementary table 3. Multivariate Cox regression models with adjustments to predict new onset Peripheral Arterial Disease (PAD), and all-cause mortality in the matched cohort.**

* for p≤ 0.05, ** for p ≤ 0.01, *** for p ≤ 0.001; HR: hazard ratio; CI: confidence interval; SGLT2I: sodium glucose cotransporter-2 inhibitor; DPP4I: dipeptidyl peptidase-4 inhibitor.

**Model 1** adjusted for significant demographics.

**Model 2** adjusted for significant demographics, and past comorbidities.

**Model 3** adjusted for significant demographics, past comorbidities, duration from earliest diabetes mellitus date to initial drug exposure date, and number of prior hospitalizations.

**Model 4** adjusted for significant demographics, past comorbidities, duration of diabetes mellitus, and number of prior hospitalizations, number of anti-diabetic drugs, and non-SGLT2I/DPP4I medications.

**Model 5** adjusted for significant demographics, past comorbidities, duration of diabetes mellitus, and number of prior hospitalizations, number of anti-diabetic drugs, non-SGLT2I/DPP4I medications, abbreviated MDRD.

**Model 6** adjusted for significant demographics, past comorbidities, duration of diabetes mellitus, and number of prior hospitalizations, number of anti-diabetic drugs, non-SGLT2I/DPP4I medications, abbreviated MDRD, HbA1c, fasting glucose.

| **Characteristics** | **New onset PAD**<br>**HR [95% CI];P value** | **All-cause mortality**<br>**HR [95% CI];P value** |
|---|---|---|
| ***Model 1*** | 0.67[0.51-0.86];0.0018** | 0.50[0.46-0.55];<0.0001*** |
| ***Model 2*** | 0.72[0.62-0.83];0.0135* | 0.52[0.47-0.57];<0.0001*** |
| ***Model 3*** | 0.75[0.64-0.89];0.0447* | 0.51[0.46-0.56];<0.0001*** |
| ***Model 4*** | 0.80[0.64-0.99];0.0441* | 0.51[0.47-0.56];<0.0001*** |
| ***Model 5*** | 0.82[0.63-0.98];0.0355* | 0.72[0.65-0.81];<0.0001*** |
| ***Model 6*** | 0.85[0.67-0.98];0.0464* | 0.72[0.65-0.80];<0.0001*** |

**Supplementary Table 4. Sensitivity analyses for exposure effects of SGLT2I v.s. DPP4I on new onset Peripheral Arterial Disease (PAD), and all-cause mortality using different models.**

* for p≤ 0.05, ** for p ≤ 0.01, *** for p ≤ 0.001; SGLT2I: Sodium-glucose cotransporter-2 inhibitors; DPP4I: Dipeptidyl peptidase-4 inhibitors; HR: hazard ratio; CI: confidence interval; PS: propensity score; IPTW: inverse probability of treatment weighting, SIPTW: stable inverse probability of treatment weighting.

| **Model** | **New onset PAD**<br>**HR [95% CI];P value** | **All-cause mortality**<br>**HR [95% CI];P value** |
|---|---|---|
| Cause-specific hazard models | 0.65[0.52-0.81];0.0001*** | 0.37[0.33-0.42];<0.0001*** |
| Sub-distribution hazard models | 0.66[0.52-0.83];0.0003*** | 0.39[0.36-0.43];<0.0001*** |
| PS stratification | 0.69[0.56-0.85];0.0004*** | 0.41[0.37-0.45];<0.0001*** |
| PS with IPTW | 0.77[0.63-0.94];0.0110* | 0.45[0.41-0.49];<0.0001*** |
| PS with SIPTW | 0.80[0.64-1.00];0.0499* | 0.46[0.37-0.58];<0.0001*** |

**Supplementary Table 5. Sensitivity analysis: Three-arm analysis results using stabilized IPTW.**

* for p≤ 0.05, ** for p ≤ 0.01, *** for p ≤ 0.001; HR: hazard ratio; CI: confidence interval; SGLT2I: sodium glucose cotransporter-2 inhibitor; DPP4I: dipeptidyl peptidase-4 inhibitor.

Adjusted for demographics, past comorbidities, duration of diabetes mellitus, and number of hospitalizations, number of anti-diabetic drugs, non-SGLT2I/DPP4I medications, abbreviated MDRD, HbA1c, fasting glucose.

| **Drug treatment** | **New onset PAD**<br>**HR[95% CI];P value** | **All-cause mortality**<br>**HR[95% CI];P value** |
|---|---|---|
| SGLT2I v.s. DPP4I | 0.72[0.59-0.87];0.0007*** | 0.25[0.21-0.28];<0.0001*** |
| SGLT2I v.s. GLP1a | 0.88[0.65-1.18];0.3905 | 0.91[0.68-1.22];0.5298 |

**Supplementary Table 6. Sensitivity analysis: Excluding patients with CKD stage 4/5 (eGFR <30), peritoneal dialysis or haemodialysis in the SGLT2I v.s. DPP4I matched cohort.**

* for p≤ 0.05, ** for p ≤ 0.01, *** for p ≤ 0.001; SGLT2I: Sodium-glucose cotransporter-2 inhibitors; DPP4I: Dipeptidyl peptidase-4 inhibitors; HR: hazard ratio; CI: confidence interval.

| | **New onset PAD**<br>**HR [95% CI];P value** | **All-cause mortality**<br>**HR [95% CI];P value** |
|---|---|---|
| **SGLT2I v.s. DPP4I** | 0.80[0.64-1.00];0.0475* | 0.58[0.52-0.64];<0.0001*** |
| Dapagliflozin v.s. DPP4I | 0.88[0.63-1.22];0.4358 | 0.53[0.45-0.63];<0.0001*** |
| Empagliflozin v.s. DPP4I | 0.84[0.60-1.17];0.2994 | 0.56[0.48-0.66];<0.0001*** |
| Canagliflozin v.s. DPP4I | 0.73[0.44-1.23];0.2374 | 0.64[0.51-0.81];0.0002*** |
| Ertugliflozin v.s. DPP4I | 0.74[0.60-0.90];0.0022** | 0.47[0.43-0.51];<0.0001*** |

**Supplementary Table 7. Sensitivity analysis: Consideration of 1-year lag time effects in the SGLT2I v.s. DPP4I matched cohort.**

* for p≤ 0.05, ** for p ≤ 0.01, *** for p ≤ 0.001; SGLT2I: Sodium-glucose cotransporter-2 inhibitors; DPP4I: Dipeptidyl peptidase-4 inhibitors; HR: hazard ratio; CI: confidence interval.

| | **New onset PAD**<br>**HR [95% CI];P value** | **All-cause mortality**<br>**HR [95% CI];P value** |
|---|---|---|
| **SGLT2I v.s. DPP4I** | 0.77[0.62-0.96];0.0210* | 0.49[0.45-0.54];<0.0001*** |
| Dapagliflozin v.s. DPP4I | 0.83[0.60-1.13];0.2349 | 0.48[0.41-0.56];<0.0001*** |
| Empagliflozin v.s. DPP4I | 0.86[0.63-1.18];0.3582 | 0.48[0.41-0.55];<0.0001*** |
| Canagliflozin v.s. DPP4I | 0.69[0.42-1.12];0.1341 | 0.56[0.46-0.69];<0.0001*** |
| Ertugliflozin v.s. DPP4I | 0.72[0.60-0.87];0.0007*** | 0.39[0.36-0.42];<0.0001*** |

**Supplementary Table 8. Sensitivity analysis: Excluding patients currently on financial aids**
* For p≤ 0.05, ** for p ≤ 0.01, *** for p ≤ 0.001; SGLT2I: Sodium-glucose cotransporter-2 inhibitors; DPP4I: Dipeptidyl peptidase-4 inhibitors; HR: hazard ratio; CI: confidence interval.

| | **New onset PAD**<br>**HR [95% CI];P value** |
|---|---|
| **SGLT2I v.s. DPP4I** | 0.72[0.58-0.88];0.0020** |
| Dapagliflozin v.s. DPP4I | 0.75[0.59-0.96];0.0239* |
| Empagliflozin v.s. DPP4I | 0.84[0.59-1.20];0.3373 |
| Canagliflozin v.s. DPP4I | 0.85[0.60-1.21];0.3770 |
| Ertugliflozin v.s. DPP4I | 0.59[0.33-1.06];0.0768 |
